# Artificial Intelligence (AI) Models for Cardiovascular Disease Risk Prediction in Primary and Ambulatory Care: A Scoping Review

**DOI:** 10.1101/2025.03.21.25324379

**Authors:** Caoimhe Provost, John Broughan, Geoff McCombe, Mark O Kelly, Mark Ledwidge, Walter Cullen, Joseph Gallagher

**Affiliations:** School of Medicine, University College Dublin, Belfield, Dublin, Ireland; UCD School of Medicine, Clinical Research Centre St Vincent’s University Hospital, Dublin, Ireland

**Author notes:** **Corresponding author:** Joe Gallagher, Dept of General Practice School of Medicine University College Dublin.

**Keywords:** Cardiovascular Disease, Artificial Intelligence, Primary Healthcare, Ambulatory Healthcare, Risk Prediction Models, General Practice

## Abstract

**Background:** Mortality from cardiovascular disease (CVD) has seen a dramatic increase over the past decades, which has led to a significant increase in the development of risk prediction models. AI-based models have been proposed as a method of enhancing traditional risk models. This review aims to describe the present state of AI risk prediction models for cardiovascular disease in primary and ambulatory care research, and in particular to determine: the stage of development these models have reached, the AI approaches used, and identifying possible sources of bias or limitations in the AI models.

**Methods:** Using the Arksey and O’Malley scoping review method, this review searched Pubmed, EBSCOHost, and Web of Science databases between 2019 and 2024, and relevant studies were identified. Data extraction was performed on eligible included studies.

**Results:** 22,860 studies were screened, and 25 articles were identified. There was a lack of external validation (20% of models) and lack of clinical impact studies (0%) in this review. A variety of AI techniques were used. Both data and algorithmic biases were commonly identified. There was a lack of geographic variation in datasets (60% were based in the USA) and only 32% of studies reported race and ethnicity data. There was poor predictor and outcome standardization. Calibration was only reported in 24% of models.

**Conclusion:** Findings from this review highlight the lack of clinical impact studies and risk of bias in current AI based models. It provides evidence for future refinement and development of AI risk prediction models in cardiovascular disease.

**Author summary:** Cardiovascular disease (CVD) is a leading cause of mortality worldwide. In this paper, we undertake a scoping review of artificial intelligence (AI) models for predicting CVD risk in primary care (PC) and ambulatory care (AC) settings.

Our findings underscore the critical role of AI in improving the prediction and prevention of cardiovascular diseases, while also highlighting the existing gaps and challenges in the application and validation of these models in primary and ambulatory care. We identified major biases in data from which models were developed, such as geographic discrepancies and inconsistencies among CVD predictors. Additionally, biases in the algorithm were significant issues that need to be addressed in the future.

We emphasize the need for multi-centered, standardized, and externally validated models to ensure their utility and reliability in clinical settings. Our research underscores the importance of addressing both data and algorithmic biases, and the necessity of external validation and clinical trials to enhance the accuracy and clinical utility of these AI models in real-world settings.

## Background

Mortality from cardiovascular disease (CVD) has seen a dramatic increase over the past few decades, rising from 12.1 million to 18.6 million cases worldwide from 1999 to 2019.^1^ In the EU, CVD is the cause of 42% of deaths, which costs the EU approximately 192 million euros per year as per 2008.^2^ Risk prediction models are used by clinicians to identify those most at risk of developing cardiovascular disease and to decide who may require interventions such as medications.

### Risk prediction models

Risk prediction models are defined as “statistical models that combine information from several markers”.^3^ ^(p.6)^ Risk prediction models produce a prediction of a patient’s risk based on the information inputted into the model.^3^ A model and its output is only reliable and clinically useful as the quality or quantity of markers (also known as risk factors) put into the model. Common uses of risk prediction models prediction of sepsis^4^, acute kidney injury,^5^ or CVD, such as coronary artery disease or heart failure.^6^ However, there are some key issues that are associated with these models, such as overly optimistic performance metrics (which may lead to models that do not generalise well on the introduction of new data)^7^ or the proper communication of the risk reliability and variability produced by the clinical prediction models.^8^ Models may not perform as well in certain populations. For example age is typically a factor that is weighed heavily, which may cause younger patients may be more likely to receive an inaccurate or diminished risk prediction value.^6^

The process of the development of a risk prediction model consists of nine main stages as outlined by Hassan et al^9^ . Steps one and two consist of generating an objective for the model, as well as defining the parameters and risk factors included. Step three is when the researchers chose a data set in which to derive their model from. This may be from a singular clinic, a series of several clinics, or a national/international database with data on the risk factors intended to be used. Step four consists of the derivation, or the creation of the model, which can use deep learning and/or machine learning techniques, and should be collaborated on with a programming specialist^9^ (to ensure AI governance, which refers to the legality and ethicality of AI in our lives^10^). Step five concerns the validation of the model, both from the dataset it was derived from (internal validation), and datasets external to the set the model was derived from (external validation). The sixth step regards the interpretation of the model, or presentation of the outcome processed by the AI model. Any underlying assumptions of the model should be identified and addressed, such as bias. Steps seven and eight regard the licensing of the model as well as the maintenance, such as updating models with new information available (i.e. newly identified risk factors or populations). The final step involved the clinical assessment of the risk prediction models. This may include “the accuracy of the model predictions, physician and patient understanding and use of these probabilities, expected effectiveness of subsequent actions or interventions, and adherence to these”.^9^ ^(p. 5)^ Complete adherence and completion of this nine stage process is essential to the proper development of a risk prediction model, as to ensure the smallest margin for error in the model programming and use.

### Artificial Intelligence in Healthcare

AI has the potential to develop new and more accurate models. Artificial intelligence is defined as a machine-based system that is designed to operate with varying levels of autonomy and that may exhibit adaptiveness after deployment, and that, for explicit or implicit objectives, infers, from the input it receives, how to generate outputs such as predictions, content, recommendations, or decisions that can influence physical or virtual environments.^8^ These modern risk prediction models have been shown to play a significant role in healthcare and risk prevention as a whole.^11^ These models have also shown that AI based models enhance clinical prediction in several domains, such as prognosis, risk assessment, and mortality prediction, compared to traditional statistics models.^12^

### Cardiovascular Disease

There are many different risk factors for CVD, and they can differ significantly between different AI risk prediction models. AI models now are encompassing the evaluation of more wide-reaching and diverse set of risk factors. These may include data collected from wearable devices (despite their inferior performance compared to other data collection forms)^13^ or CVD risk in conjunction with other disease diagnoses such as cancer or diabetes.^14, 15^ However, it is important to mention that AI models can be imperfect, as issues may arise such as bias (or systematic errors, such as in model choice, data used to train the model, or model evaluation, present in model predictions^16^), accountability (“a set of protocols to evaluate the conduct of an entity, as well as an obligation to report or justify one’s actions, specially if these may result in any wrongdoing” ^17^ ^(p. 3)^), and transparency (ensuing information about the derivation and development of the model is accessible to the public^18^). The presence or lack of these issues can lead either to a more or less accurate AI prediction model, and these issues may have a real-life impact on patient treatment and outcome.

Various CVD AI risk prediction models do exist currently in the PC and AC setting. These models use various different tests routinely available in GP or ambulatory care settings with the goal of predicting cardiovascular disease or events or death. For example, results from routine blood tests, such as total cholesterol levels of serum creatinine levels,^19^ as well as electrocardiogram results ^20^ may be used in order to derive these AI models.

### Gap in the Literature

In current literature, there exists a gap in the knowledge consolidated and reviewing AI risk prediction models evaluating CVD in primary care (PC) and ambulatory care (AC). Cai et al. 2021 reviewed CVD AI risk prediction models and their validation, concluding that there was larger focus on developing new models than validating (or evaluating model performance/accuracy) existing ones^21^, but there is a notable lack of a focus on PC or AC accessible models. Silva et al. reported that errors in the AI models such as explainability, bias or data leakage need to be properly monitored and heavily analysed to assure both ethical and legal AI usage^22^, which has not been analysed in pre-existing PC or AC literature.

Therefore, this paper aims to address three main aims:

1. To determine what stage of development these models have reached.
2. To determine what AI approaches have been used in the derivation and validation of these models.
3. To identify possible sources of bias or limitations in the AI models identified.

## Methods

This study chose to conduct this review using a scoping review methodology, as it aligns with the objective of gaining a complete and overarching review of the clinical value risk prediction models involving AI for CVD for primary care. Scoping reviews, which are defined as “knowledge syntheses used to collect, evaluate, and present findings from existing research on a topic” ^23^ ^(para. 5)^ are fundamental to the furthering and summarising existing knowledge from around literature and establishing gaps. This study followed the six-stage PRISMA scoping review method outlined by Arksey and O’Malley’s ^24^, as well as Levac et al.^25^

### Stage 1: Identifying the research question

With regard to both the gap in the previous knowledge surrounding CVD AI risk prediction models used in primary/ambulatory care settings and the objectives of this study, the research question was developed:

### What does the pre-existing literature report on artificial intelligence cardiovascular disease risk prediction in PC and AC, specifically: what stage of development these models have reached, what AI approaches have been used in the derivation and validation of these models, and what possible sources of bias or limitations in the AI models identified?

### Stage 2: Identifying relevant studies

An initial search of databases including PubMed/Medline, Web of Science, and EBSCOHost was taken on 01/06/2024. To search these databases, a series of medical subject heading (MeSH) terms were identified in alignment with this study’s research question. These terms were then grouped together based on their subject matter, and so that resulting searches would include at least one search term of the four groups included (see **appendix 1**).

### Stage 3: Selecting studies

Following the consolidation of the identified studies from the databases, duplicates were removed. Two reviewers read the identified studies titles and abstracts, and full texts were retrieved for all the studies. Studies found to be relevant to the study had a full-text review conducted by one researcher. This process is outlined in **Figure 1** below

**Fig 1.**
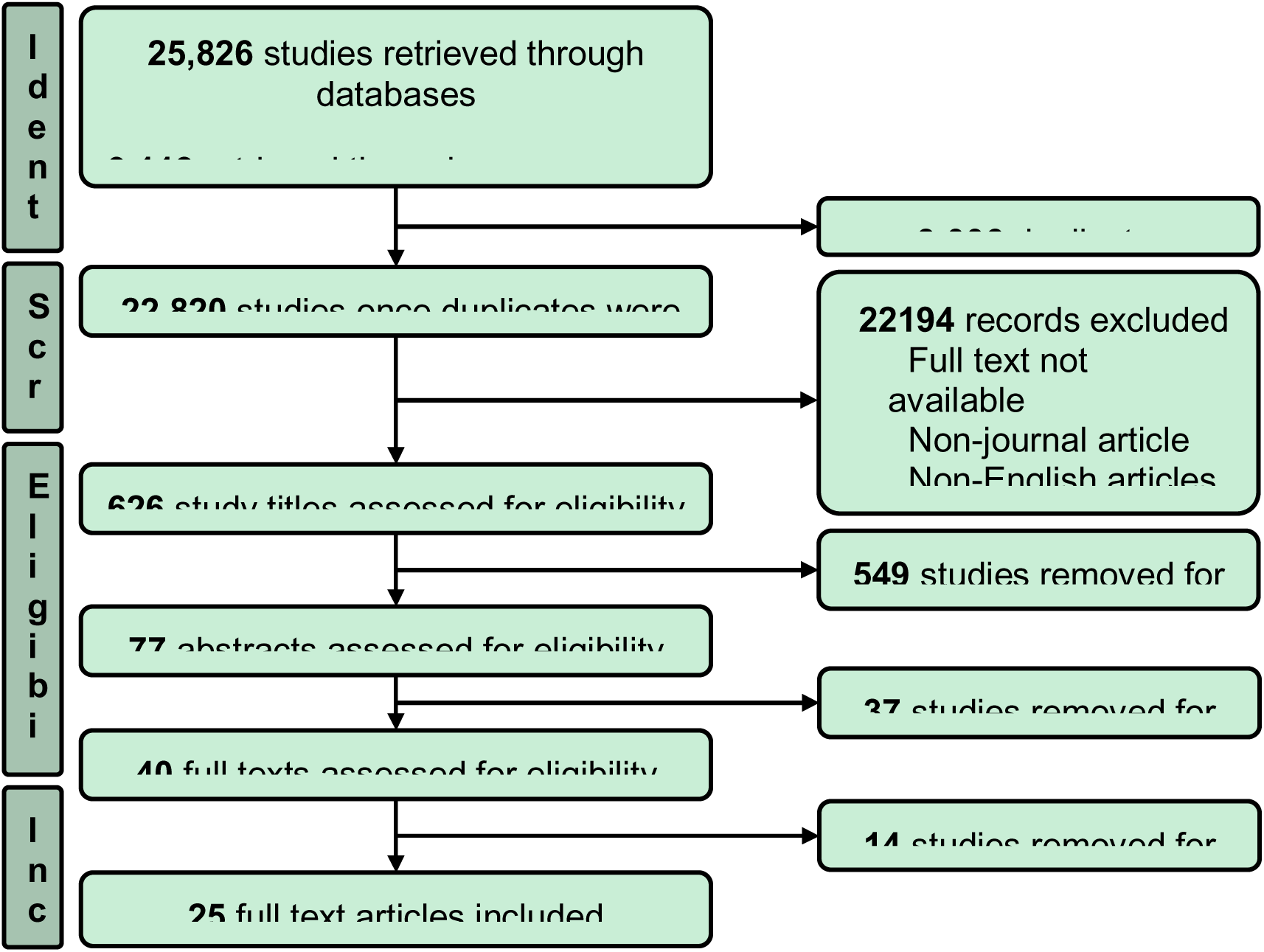
PRISMA flow chart of this scoping review methodology

Eligibility criteria were developed for this scoping review. Included studies were published between 2021 and 2024 and involved adult human participants. These studies also must include 1) a multivariable AI model relating to the risk prediction of CVD, 2) an outcome measure of either CVD or cardiovascular events or death and 3) conducted in a hospital outpatient or primary care setting.

Excluded studies for this review included non-English papers, those studying paediatric patients only, as well as 1) studies based on single predictors only, 2) clinical prediction rules developed from patients with acute cardiovascular events or cardiac surgery only or hospital inpatients only, and 3) models involving tools not available routinely in primary care (see **appendix 2**).

### Stage 4: Charting the data

After analysing all 25 full texts, a data extraction table was created. This included categories such as: Author and year of publication, location of study and datasets used, outcome and output of the risk prediction model, results of the AI risk prediction model incusing mentions of discrimination or calibration, the type of AI used, predictors used model development, limitations of the study, and the models most recent stage of validation. A copy of the table created is shown in **Table 1**.

**Table 1:** The worldwide distribution of studies included in this review.

| Country name | Number of studies based in the country |
| --- | --- |
| United States of America | 15 |
| China | 7 |
| South Korea | 4 |
| Brazil | 1 |
| Taiwan | 1 |
| Austria | 1 |
| Japan | 1 |
| Canada | 1 |

### Stage 5: Collating, summarising, and reporting results

This review focused primarily on emphasising key elements of the research found during stage 4 of the PRISMA methodology. In accordance with Tripod AI guidelines^26^ as well as consultation from experts in the field of AI risk prediction models and primary/ambulatory care, six overarching result categories were described.

### Stage 6: Consultation

This paper underwent a consultation exercise by two individuals in order to enhance its usefulness to policymakers, practitioners and other readers.^24^

Upon this consultation, no further studies which fit the inclusion criteria were brought to the attention of the authors. However, one individual noted that there was a lack of the genetic and biomarker data such as natriuretic peptide data used in datasets to train the AI model, despite its growing popularity as a method of risk prediction for CVD. This clinical data would provide further information into the genetic predispositions, including epigenetic factors which may increase, decrease, or have a neutral effect on the likelihood of a patient to develop CVD.^27^ It was also highlighted regarding the need to ensure adequate supervision of the models in clinical settings. Supervision in machine learning refers to the process of training a model using a labelled dataset, where the input data is paired with corresponding outputs (labels). This process allows the model to learn the relationship between inputs and outputs, enabling it to make predictions or decisions when presented with new, unseen data. This is important in clinical practice to ensure appropriate input data is utilised.^28^

## Results

### Study population and statistics

In total, 25 full text articles were included in this review, all which were published between 2021 and 2024. These studies contained a combined 32 unique datasets from four distinct regions (North America, South America, Asia, and Europe), as shown in **Figure 2** and **Table 1** below.

**Fig 2.**
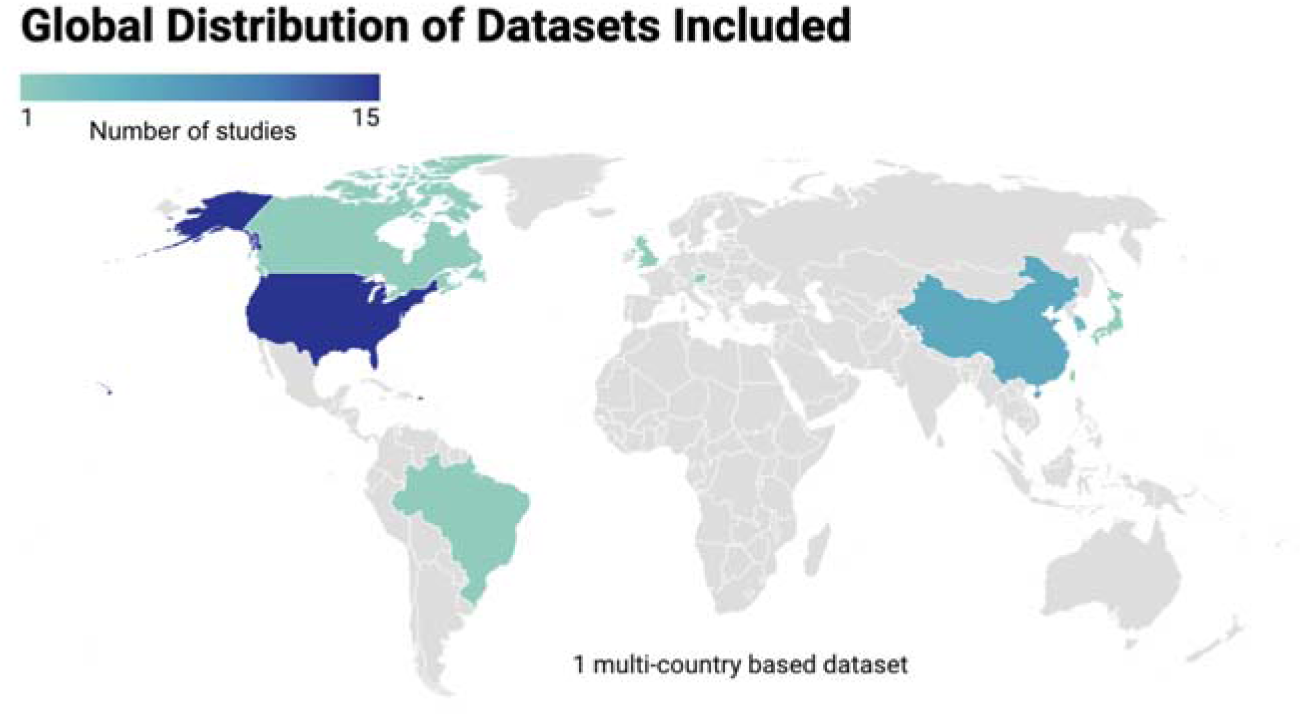
The worldwide distribution of datasets included in this review

In the 25 studies included, the average age of participants was approximately 62.7 years old (41.2 ^29^-78.8 years ^30^). 52.1% of included participants were male, and of the 23 studies that did specify the sex ratios, 16 contained a cohort with a roughly balanced proportion of male to female participants (between 40% - 60% male). Eight studies included race and/or ethnicity statistics of their cohort.^31, 32, 33, 34, 35, 36, 37, 38^ 59.8% of participants were White or European American, with the African American/Black/Black British population making up about 23.0% of all cohorts.

While 21 included studies focused generally on the current population, five studies concerned sub-groups of the population, rather than the general public. These concerned oncology patients (n=1, 4.8%),^39^ women and women’s health (n=1, 4.8%),^37^ older patients (n=2, 9.6%),^30, 40^ prediabetes or diabetes outpatients (n=1, 4.8%),^40^ and African-American patients (n=1, 4.8%).^41^

The average sample size of all the studies was approximately 89783 participants (289 ^42^ −691645 participants^43^). The distribution of study sizes is shown in **Figure 3**. A total of four different study settings were identified, which included an in person primary care setting, hospital (inpatient or outpatient) setting, online or telehealth surveys, and referral centre setting as shown in **Figure 4**.

**Fig 3.**
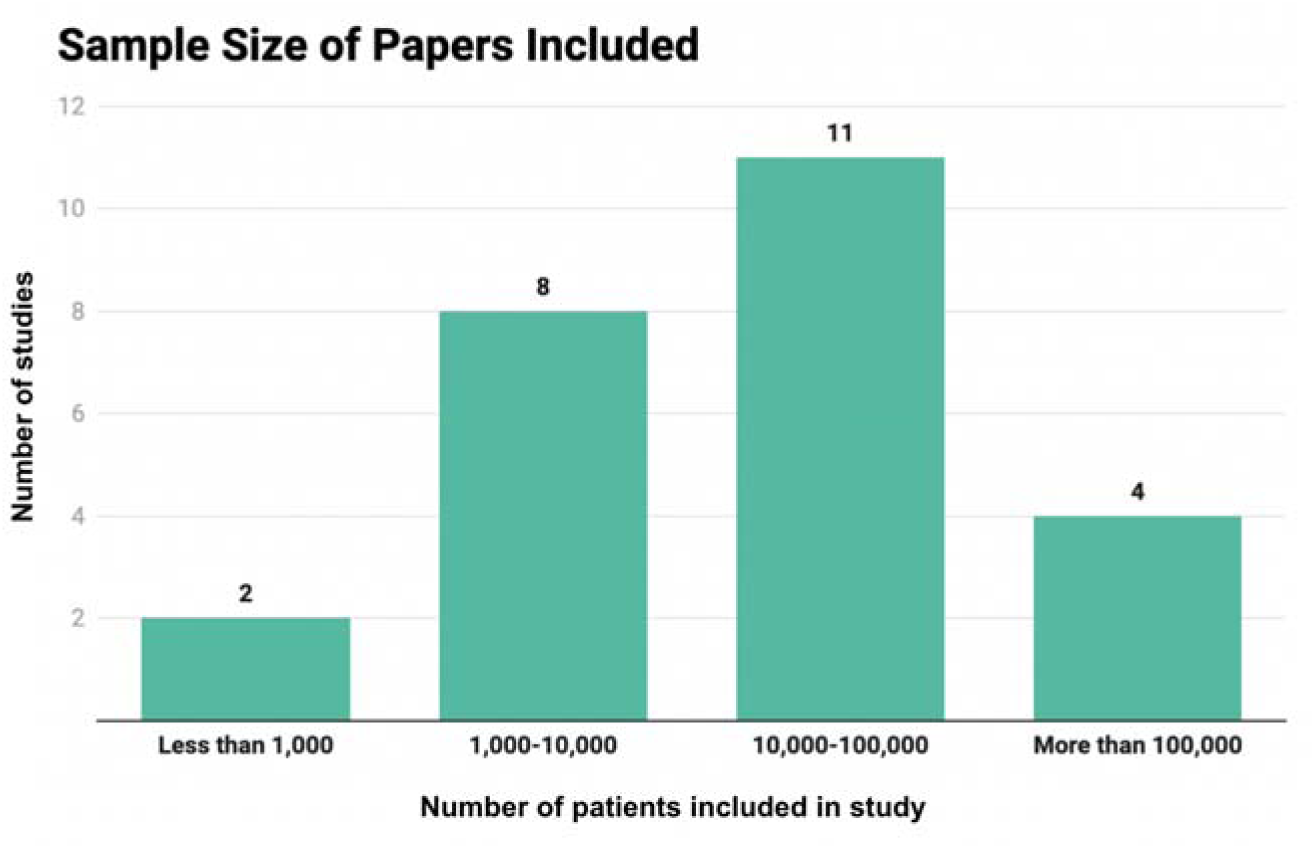
Patient sample size for included studies

**Fig 4.**
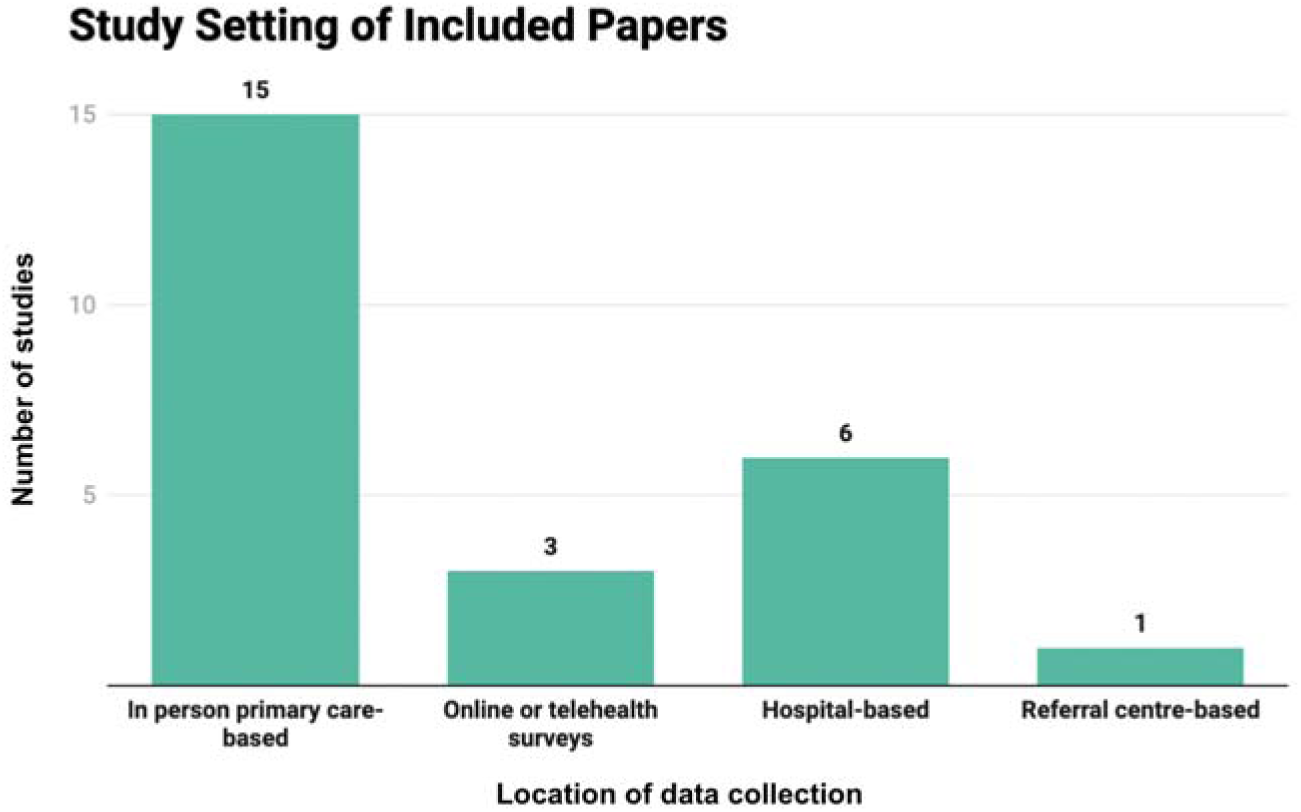
Setting of data collection for included studies

### Setting of data collection

A significant majority of the included texts (n=15) were conducted in a community or clinical care setting, followed by those in a hospital or inpatient setting (that used tools available to PC or AC practitioners) (n=6),^30, 32, 37, 44, 45, 46^ those in a virtual or online survey (n=2),^42, 47^ and studies conducted in a referral centre (n=1).^48^ Of the 25 full-texts included, 11 specified that only completed data (i.e. patients with complete electronic health records and/or electrocardiograms) were included in the training or validation sets for the AI model. The minority of included studies used multiple databases (n=8) for the training (and the validation) of their risk prediction model ^29, 36, 37, 38, 45, 46, 48, 49^ with the rest relying on one database in order to train, test, and validate the models

### Use of CVD predictors

Across all models, 255 unique parameters used by the included AI risk prediction CVD models were identified. Age (n=18), body mass index (BMI) (n=13), and history of or current habit of smoking (n=10) were the most common parameters used in the models. Of the 255 parameters, 57 were found to have been included in models from two or more studies, and 27 in three or more papers, which are shown in **Figure 5**. It is also important to highlight the presence of vague predictors, such as “12-lead raw electrocardiogram (ECG) data”,^50^ but not the elements they extracted from that specific data. This may be an example of unsupervised training.

**Fig 5.**
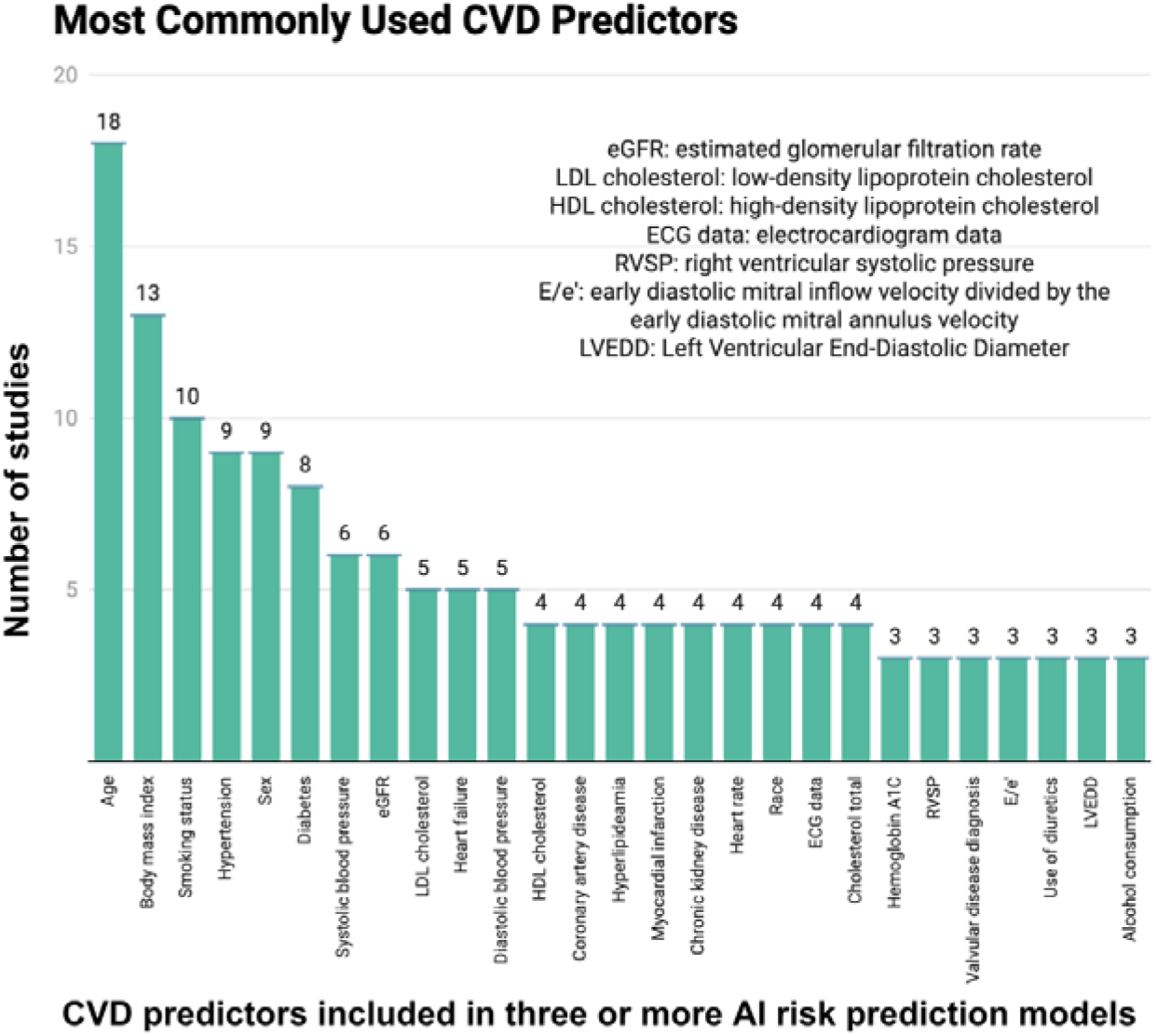
The most used CVD predictors present in three or more included AI risk prediction models

### CVD outcome measurement and method

All included texts in this review stated what their AI model was predicting by the patient outcome. The predictions of an unspecified CVD diagnosis were found to be the most common (n=6).^29, 39, 41, 43, 44, 47^ Other predictions included the onset or diagnosis of atrial fibrillation (n=5),^30, 33, 35, 42, 46^ the development of heart failure (n=4),^31, 36, 40, 47^ prediction of mortality (with all but one describing predictions of all-cause mortality) (n=4),^32, 44, 45, 48^ “net adverse clinical events” such as all-cause death, myocardial infarction, stroke or major bleeding (n=3),^51, 52, 53^ coronary artery disease events (n=2),^34, 38^ and aortic regurgitation development (n=1).^54^ One of the models had a composite endpoint -prediction of mortality and/or cardiovascular events.^44^ 15 models mentioned periods of time their prediction was for, with the most common being exactly 1-year following the initial data collection (n=7),^32, 34, 35, 43, 48, 52, 54^ as shown in **Figure 6**.

**Fig 6.**
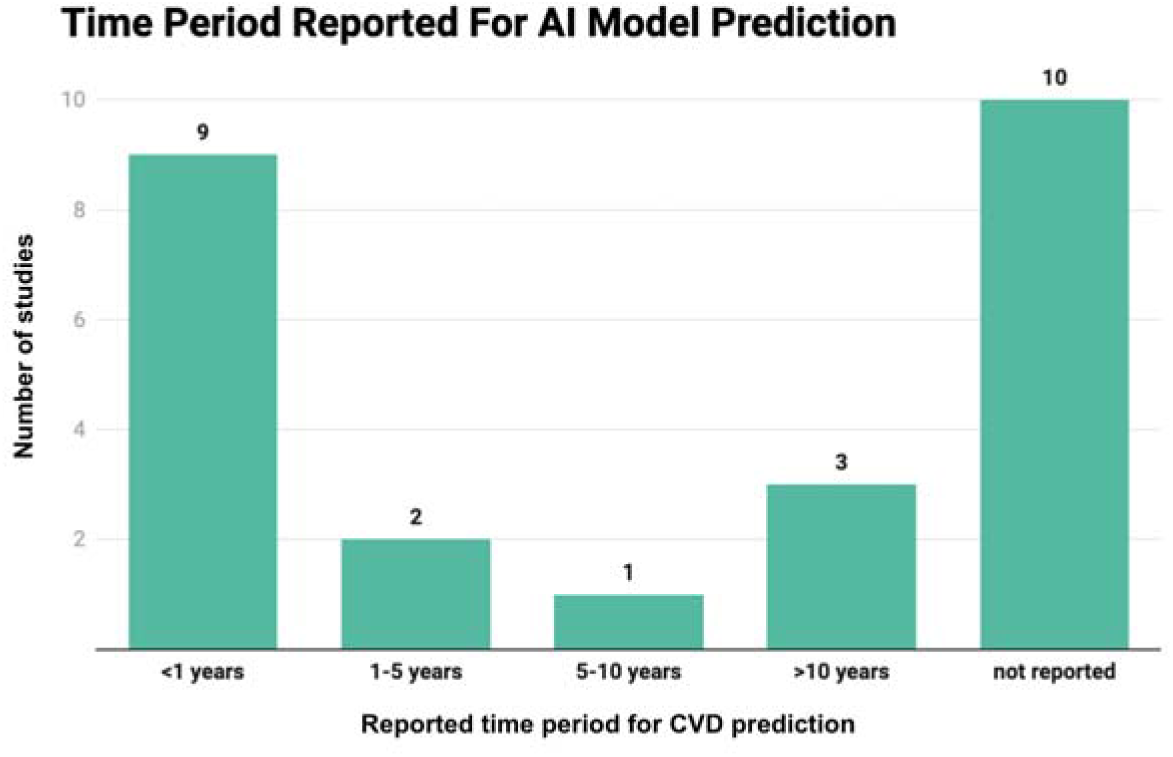
Reported time period for AI model prediction for CVD

### AI model development method

In AI, there exists two types of learning: machine (which involves “the capacity of systems to learn from problem-specific training data to automate the process of analytical model building and solve associated tasks” ^55^ ^p.1^), and deep learning (which uses artificial neural networks to emulate the human brain and its process of learning).^55^ An overwhelming majority of studies in this review developed machine learning models (n=19) rather than deep learning models^31, 35, 41, 42, 44, 46^, as shown in **Figure 7**.

**Fig 7.**
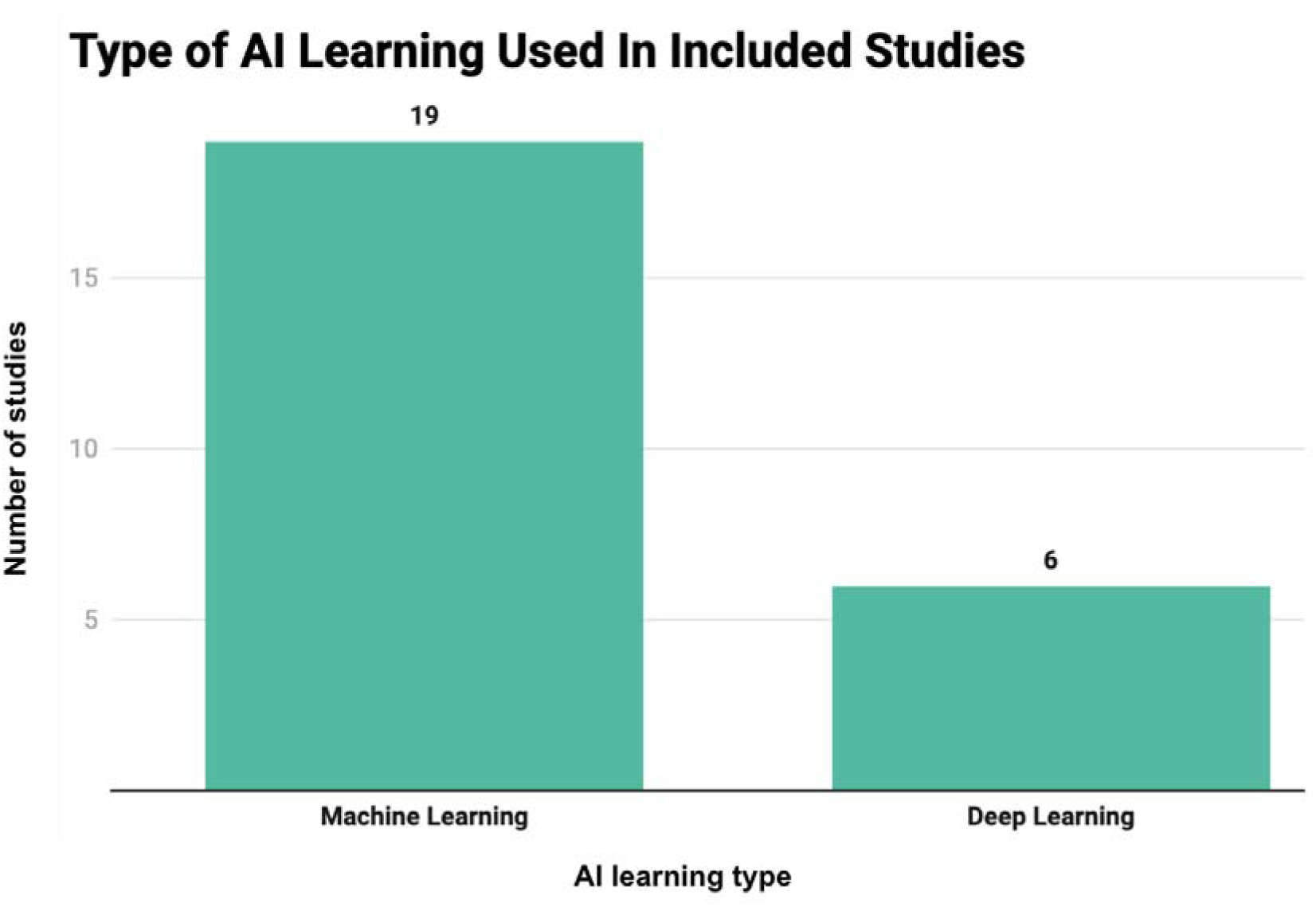
The form of AI learning used in studies

A variety of AI techniques were used for model development: random survival forest (n=12), proportional hazard technique (n=11) and artificial neural networks (n=7)^31, 35, 39, 41, 42, 44, 46^ were the most common. Other techniques included gradient-boosting decision trees (n=6),^34, 36, 40, 48, 49, 52^ support vector machines (n=4)^29, 30, 34, 40^ and Naive Bayes classifiers (n=1).^43^

The most common measure of model discrimination, or the process undertaken to identify the most accurate or useful AI model from a set of competing developed models,^56^ was a form of an AUC (area under the curve) measure (n=18). Confidence intervals were also recorded in 10 studies. Specificity and sensitivity are measures which evaluated the number of true versus false negatives (specificity) or positives (sensitivity) that are reported by the AI model.^57^ Sensitivity was a measure mentioned in seven included model evaluations,^33, 34, 35, 38, 42, 43, 46^ while specificity measures were present in eight studies.^29, 30, 33, 34, 35, 38, 42, 46^ Calibration, or the comparison between predicted and actual results of patients, was only discussed in a minority of studies (n=6),^36, 40, 47, 48, 51, 53^ despite its importance in model development.

### Study limitations and model validation

The most commonly mentioned limitations were limitations with the methodology or development of the model, which were mentioned in 17 papers. These were then followed by limitations in AI model validation (n=14), dataset limitations (n=11), as well as limitations with the AI or technological aspects of the models (n=6).^30, 35, 43, 44, 52, 53^ Specifically on model validation, it was found that the majority of models included had reached an internal validation state (n=18, 72%). While a few papers were able to conduct external validations (n=5),^30, 34, 36, 48, 49^ there were no cases in which a clinical trial was conducted to evaluate the effectiveness of these models, as shown in **Figure 8**.

**Fig 8.**
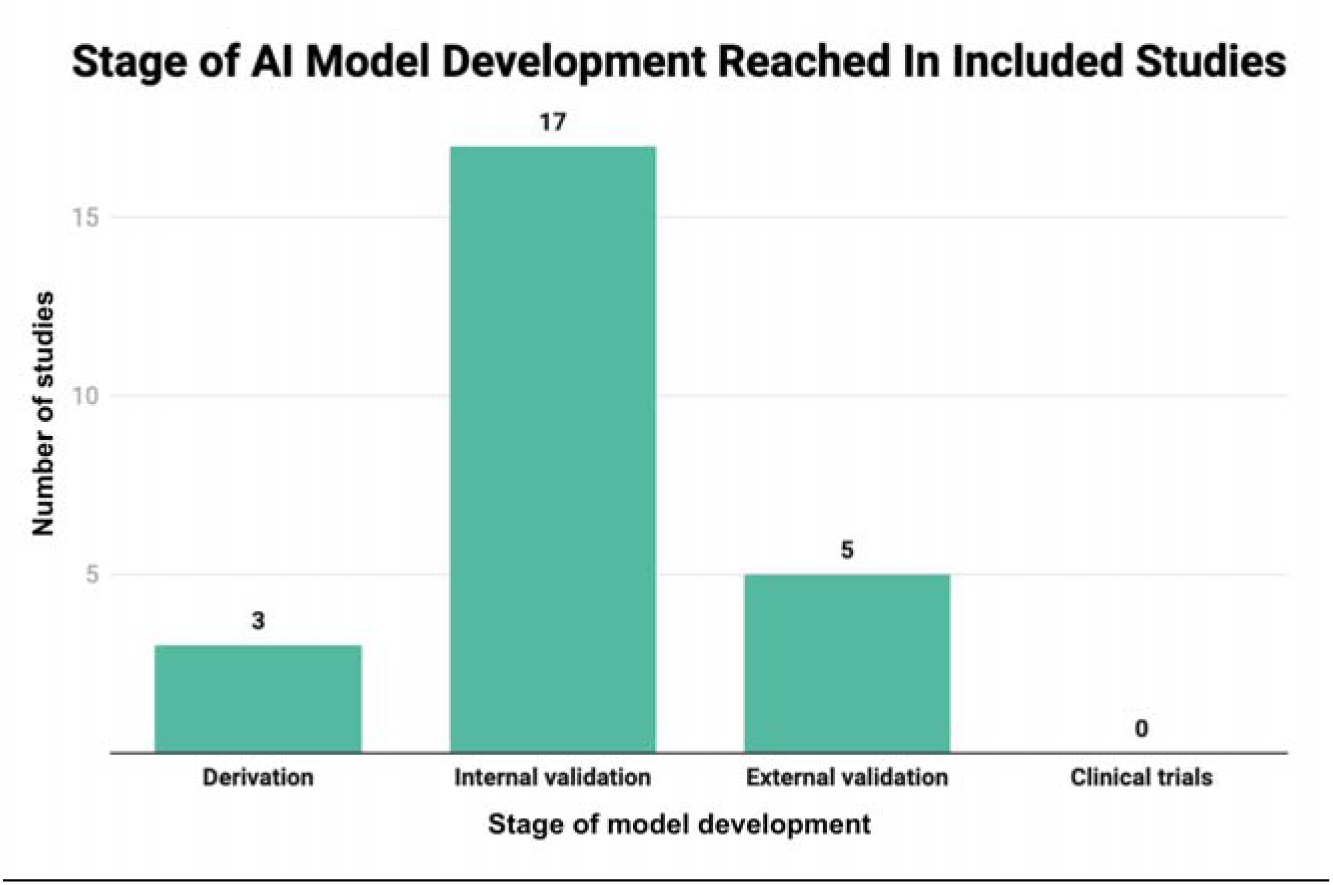
Graph of the AI model stage of development reached by studies in this review

## Discussion

Since 2021, the environment surrounding AI risk models has grown substantially and more accessible to the average researcher or clinician due to the increasing presence of AI practices in daily procedures.^58^ Overall, the aim was to 1) assess the current stage of development these models have achieved, 2) to identify the AI techniques used in the creation and validation of the models, and 3) to pinpoint potential sources of bias or limitations in the identified AI models. Findings of this study identified the overall trends with model development, specifically the data and algorithmic biases present in the models included, as described below.

### Major data bias findings

Data biases, or bias resulting from the presence of unfair or unrepresentative data that helped develop the AI model,^59^ may lead to inaccurate CVD predictions or biased predictions associated with certain population groups.^60^

#### Geographic discrepancy

Through the population analysis of the included studies, an overwhelming majority of databases were based in the United States of America or in Eastern Asia (i.e. China, Japan, Taiwan, Korea, etc.). However, there were no datasets used from the African or Oceania or Middle Eastern regions, highlighting a significant gap in the knowledge. This finding has been mentioned in previous reviews of CVD AI risk models.^21, 50^ Additionally, several studies did not provide statistics regarding the age, sex, or racial distribution of their study cohort, not fully explaining any underlying biases or over/under presence of certain populations. This discrepancy in data usage and provision from different geographic regions shows the importance of using multi centred studies should be encouraged.

#### Consistency among CVD predictors and predictions

Overall, age was found to be the most commonly used predictor of CVD or related events, having been mentioned in models from 18 papers. BMI, having a history of smoking/or being a current smoker, as well as a history of hypertension and the patient’s sex were also found to be parameters that were significant in predictions, due to their overwhelming use in nine or more studies. Standardisation, or the technical specifications necessary for AI (and some other technologies) to perform effectively, was highlighted by Golenkov et al ^61^ as a novel difficulty for AI in its staggering process. This is as standardisation (or lack thereof) may impact the compatibility and interoperability between models or different standards.^50^ Creating a “gold standard”, or a commonly agreed upon method of measurement, or measure where a person is categorised as having developed a disease is important, as it allows for two AI models to be used and predict a similar outcome, even if they use different types of learning or parameters.^60^

Similarly, the most common outcome prediction of the models was an unspecified CVD diagnosis category. While specific cardiovascular diseases, such as heart failure or atrial fibrillation are acceptable outcomes for these models, CVD, which is a “collective term designating all types of affliction affecting the blood circulatory system, including the heart and vasculature”,^62^ ^p.1^ is not specific enough to be able to understand what exactly is being predicted by the model.

### Major algorithmic bias findings

On the other hand, algorithmic biases, or biases introduced by the AI algorithms themselves, may also lead to inaccurate CVD predictions or the impression of AI models to make predictions treating groups fairly.^63^

#### Model development and its impact on clinical practice

The results of this study found a lack of development of AI risk prediction CVD models, especially in relation to independent external validation. The goal of the majority of the included studies is to aid in the individualised diagnosis or risk calculation of a patient in regards to CVD, and this can only be properly achieved on a worldwide basis if there is a focus on (specifically external) model validation.^64^ This overall trend of an large number of internally validated sources may be due to a focus by researchers on the development of new and diverse models, rather than the testing of previously designed risk predictor models in environments or with patients previously unseen or unused during the models derivation or internal validation, such as validation the model among a population of a different background or ethnicity. According to the six-stage process as outlined by de Hond et al. (2022)^50^, 20 of the included models in this review have only reached stage two: Development of the AI prediction model (which includes both derivation and internal validation).^50^ It is also important to note that no model surpasses stage three: Validation of the AI prediction model, as no study was found to have undergone clinical trial to not only test for the accuracy of the model, but also its usefulness, such as with patient or doctor-AI interactions, addressing AI security risks, and impact assessment of the model in clinical practice.^50^

Model discrimination, which are the biases and discriminatory outcomes that arise when AI risk prediction models make predictions or decisions,^56^ was found to have been investigated in 92% of included studies. Discrimination in model performance or predictions made, whether explainable or unexplainable, may arise from bias in the data and/or the algorithm,^65^ and may have both legal and ethical considerations, such as the neglect of certain ethical groups or geographic locations.^21^ Model calibration measures is the process of adjusting the predicted probabilities of a model so that they accurately represent the true likelihood of an event,^66^ was conducted in only 24% of models. Conducting these calibration tests is crucial to providing accurate risk predictions to patients, as to avoid providing incorrect real-life medical interventions to patients.^26, 67^ Due to the emerging environment around AI in healthcare decision-making, it is crucial that these models undergo both discrimination and calibration checks, as to ensure these systems are both fair and trustworthy.^68, 69^

### Implications of AI risk prediction models for CVD in PC or AC

This research underscores the critical role of AI in improving the prediction and prevention of cardiovascular diseases, while also highlighting the existing gaps and challenges in the application and validation of these models in primary and ambulatory care.

A focus regarding indemnifying gaps in the pre-existing literature, specifically regarding the presence of algorithmic and data biases, primarily on the importance of external validation and clinical trials, as in stark contrast to the current trend of prioritizing only the development and internal validation of these models. Clinical researchers may be able to externally validate models, such as ones included in this review or newly created models, as to aid in discovering a model that is accurate as well as clinically relevant and useful for primary or ambulatory care clinicians. This also applies to global research funding bodies, which should be focusing on funding projects taking the step to externally validate new or existing models, as well as creating a standardisation of models and their parameters, and/or global datasets used, rather than solely promoting the development of novel AI models following in the steps of previous study methodology.

### Strengths & limitations

#### Study strengths

The use of a scoping review methodology was a significant strength of this article, due to the rigorous screening and assessment of included studies, which was conducted using the Arksey and O’Malley scoping review framework.^24^ This study was rigorous in its adherence to the method, ensuring all steps from the development of the research question to the consultation of the findings with other experts in the field were thoroughly followed.

#### Study limitations

There are several limitations to this review, including a limit to only including full text, accessible, English language studies. This may have led to an incomplete scope of the existing literature or models that have been researched related to this review, and may have impacted the findings of this study, such as the global distribution of datasets used. As AI risk prediction models is a rapidly evolving field, conducting this review even a month later, may have also produced different results, such as additional models.

As well, this review did not aim to summarize the clinical applicability or effectiveness of the models, especially compared to traditional or non AI risk prediction models. While it is important to understand the general structure and key aspects of these models (such a time to events or parameters included), their usefulness is a very important investigation, as to continue to improve upon the models, and the future of AI in healthcare in general. This makes the comparison between AI and non-AI models an essential area for further research.

Within the 25 included studies, none used the precision recall AUC, which is a useful tool in model discrimination, in which there is a focus on achieving a high number of true positives (where the model was correct in its prediction for a given patient). AUC-ROC tends to be a more “simplistic” evaluation of a risk prediction model (though still useful in the validation of the model), whereas a precision recall AUC may be more beneficial in cases where the true-positive and true–negative outcome are not balanced in their costs (development of CVD or not).^70^ This highlights the need to define measures of model accuracy that can be used across studies. Finally, in the scoping review methodology, there is no formal evaluation of the quality of the included studies as is typical in systematic reviews.

## Conclusion

In conclusion, there are a significant number of studies involved in the development and internal validation of novel and diverse AI risk prediction models for CVD in a PC or AC environment. This scoping review highlights that there needs to be a multi centred, standardisation, and external validation focus to these models going forward, as to continue to discover the clinical utility of risk prediction models using AI. Gaps identified by authors will act as future guidance for PC and AC researchers, research funding avenues, and governmental policies regarding AI usage and development.

## Data Availability

All data produced in the present work are contained in the manuscript

## Acknowledgements

Thank you to the University College Dublin School of Medicine for generously funding this research through the AI in Medicine Scholarship and for facilitating its development through the Student Summer Research Award (SSRA) program.

## Appendix 1

(Heart failure [MeSH Terms] OR Atrial fibrillation [MeSH Terms] OR Acute coronary syndrome [MeSH Terms] OR Cardiovascular diseases [MeSH Terms] OR Myocardial infarction [MeSH Terms] OR coronary Artery disease [MeSH Terms] OR stroke [MeSH Terms] OR cerebrovascular disorders [MeSH Terms] OR heart disease [MeSH Terms] OR arrhythmias, Cardiac [MeSH Terms] OR Angina, Pectoris [MeSH Terms] OR myocardial ischemia [MeSH Terms] OR All-cause mortality [MeSH Terms] OR Coronary artery bypass [MeSH Terms] OR Percutaneous transluminal coronary angioplasty [MeSH Terms] OR Heart failure [Text Word] OR Atrial fibrillation [Text Word] OR Acute coronary syndrome [Text Word] OR Cardiovascular diseases [Text Word] OR Myocardial infarction[Text Word] OR coronary Artery disease [Text Word] OR stroke [Text Word] OR cerebrovascular disorders [Text Word] OR heart disease [Text Word] OR arrhythmias, Cardiac [Text Word] OR Angina, Pectoris [Text Word] OR myocardial ischemia [Text Word] OR All-cause mortality [Text Word] OR Coronary artery bypass [Text Word] OR Percutaneous transluminal coronary angioplasty [Text Word])

AND

(Machine learning [MeSH Terms] OR Deep learning [MeSH Terms] OR Artificial intelligence [MeSH Terms] OR Unsupervised [Text Word] OR Supervised [Text Word] OR Deep learning [Text word] OR Machine learning [Text Word] OR Artificial intelligence [Text Word])

AND

(Risk [MeSH Terms] OR Mortality [MeSH Terms] OR Risk prediction [Title/Abstract] OR Prediction [Text Word])

## Appendix 2

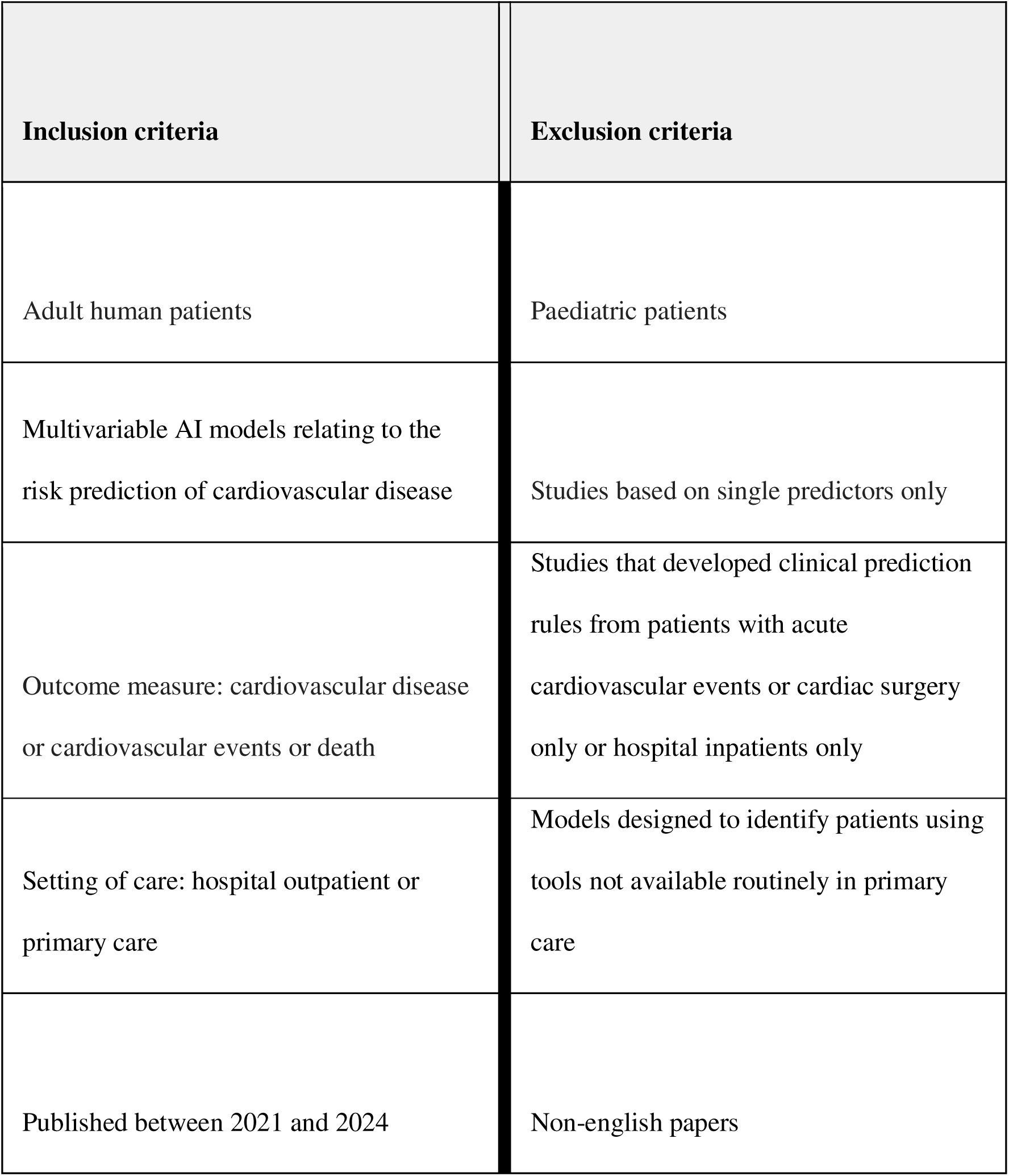

| Inclusion criteria | Exclusion criteria |
| --- | --- |
| Adult human patients | Paediatric patients |
| Multivariable AI models relating to the risk prediction of cardiovascular disease | Studies based on single predictors only |
| Outcome measure: cardiovascular disease or cardiovascular events or death | Studies that developed clinical prediction rules from patients with acute cardiovascular events or cardiac surgery only or hospital inpatients only |
| Setting of care: hospital outpatient or primary care | Models designed to identify patients using tools not available routinely in primary care |
| Published between 2021 and 2024 | Non-english papers |

